# REDDI: A Riemannian Ensemble Learning Framework for Interpretable Differential Diagnosis of Neurodegenerative Diseases

**DOI:** 10.64898/2026.04.10.26350617

**Authors:** Mario Roca, Giovanni Messuti, Dmytro Klepachevskyi, Marianna Angiolelli, Simona Bonavita, Francesca Trojsi, Matteo Demuru, Emahnuel Troisi Lopez, Sylvain Chevallier, Florian Yger, Ausra Saudargienė, Pierpaolo Sorrentino, Marie-Constance Corsi

## Abstract

Neurodegenerative diseases such as Mild Cognitive Impairment (MCI), Multiple Sclerosis (MS), Parkinson’s Disease (PD), and Amyotrophic Lateral Sclerosis (ALS) are becoming more prevalent. Each of these diseases, despite its specific pathophysiological mechanisms, leads to widespread reorganization of brain activity. However, the corresponding neurophysiological signatures of these changes have been elusive. As a consequence, to date, it is not possible to effectively distinguish these diseases from neurophysiological data alone. This work uses Magnetoencephalography (MEG) resting-state data, combined with interpretable machine learning techniques, to support differential diagnosis. We expand on previous work and design a Riemannian geometry-based classification pipeline. The pipeline is fed with typical connectivity metrics, such as covariance or correlation matrices. To maintain interpretability while reducing feature dimensionality, we introduce a classifier-independent feature selection procedure that uses effect sizes derived from the Kruskal-Wallis test. The ensemble classification pipeline, called REDDI, achieved a mean balanced accuracy of 0.76 (±0.05) across five folds, representing a 9% improvement over the state of the art, while remaining clinically transparent. As such, our approach achieves reliable, interpretable, data-driven, operator-independent decision-support tools in Neurology.

## 1. Introduction

As global demographics shift toward an aging population, the prevalence of neurodegenerative diseases, including Mild Cognitive Impairment (MCI), Parkinson’s Disease (PD), Multiple Sclerosis (MS), and Amyotrophic Lateral Sclerosis (ALS), has risen dramatically. Collectively, these four conditions affect over 85.8 million individuals worldwide (71 million with MCI [1], 11.77 million with PD [2], 2.8 million with MS [3], [4], and 360,000 with ALS [4]). However, while these diseases arise from distinct pathophysiological processes, they all result in extensive reorganization of the brain structures and functional activities. The neurophysiological signatures of this reorganization, have not been clearly identified, limiting the ability to distinguish between these conditions using neurophysiological data alone, and to effectively track these changes longitudinally. In fact, most existing studies focus on binary classification tasks—distinguishing between healthy subjects and patients. In this manuscript, we ask whether neurodegeneration induces a constrained set of large-scale reorganization patterns in fast brain dynamics that are common across diseases, and if disease-specific deviations from these patterns be identified through the structured integration of whole-brain connectivity information and used for automated differential diagnosis. On the one hand, non-invasive neuroimaging, such as electroencephalography (EEG) and magnetoencephalography (MEG), offers a window into observing and characterizing fast whole-brain dynamics. On the other hand, interpretable machine learning (ML) has been developed to identify features that characterize and distinguish groups, while also quantifying the relative importance of each feature.

However, most classification studies lack interpretable pipelines (for example, performance metrics, such as the accuracy, are typically reported disregarding the relevance of the individual features). Furthermore, these features typically rely on local brain measurements, neglecting the critical role of altered inter-regional communication in neurodegenerative diseases. In a prior study [5], it was shown that a parsimonious set of phase-based, edge-centric connectivity metrics, rather than nodal or amplitude-based features, yields maximally effective and robust cross-disease discrimination (balanced accuracy: 67.1% vs. 35% chance-level). However, while these features provide complementary information, the question of how to optimally integrate them to enhance classification performance remains unresolved. A key challenge lies in the high-dimensionality of functional connectivity data, which is typically represented as adjacency matrices. Geometric methods, modelling covariance matrices on Riemannian manifolds, have emerged as a promising solution. This framework enables operations such as averaging, smoothing, interpolation, and extrapolation of matrix-valued data [6], [7], [8]. These methods have been successfully applied in Brain-Computer Interface research to robustly detect mental states, suggesting their potential for clinical diagnostic applications.

Building on the aforementioned challenges and opportunities, we hypothesize that integrating functional connectivity estimators with Riemannian geometry and ensemble learning would enhance the performance of multi-disease classification in neurodegenerative diseases. In particular, we aimed at identifying a common backbone of features (i.e., nodes or edges) allowing to distinguish among all diseases at once (as opposed to identifying disease specific features). Furthermore, we posited that the selected features could characterize the disease-specific spread on the whole-brain scale [9]. To test these hypotheses, we analyzed a large dataset of source-reconstructed MEG recordings from a diverse cohort of 109 patients, including individuals diagnosed with mild cognitive impairment, amyotrophic lateral sclerosis, Parkinson’s disease, and multiple sclerosis (MS). To analyse the data, we develop a novel framework that leverages and expands interpretable machine learning methods to support differential diagnosis while preserving clinical interpretability.

## 2. Materials and Methods

### 2.1 Data collection and signal processing

The dataset comprises 109 patients with various neurological diseases, namely Amyotrophic Lateral Sclerosis (ALS), Mild Cognitive Impairment (MCI), Parkinson’s Disease (PD), and Multiple Sclerosis (MS), recruited from the Hermitage Capodimonte Clinic in Naples and collaborating institutions. The protocol was approved by the “Comitato Etico Campania Centro” (Prot.n.93C.E./Reg. n.14-17OSS). In accordance with the Declaration of Helsinki, all participants provided written informed consent. The demographics and the source of the data are summarized in Table 1. Diagnoses of MCI, ALS, PD, and MS were established according to the National Institute on Aging–Alzheimer’s Association criteria [10], the El-Escorial criteria [11], the United Kingdom Parkinson’s Disease Brain Bank criteria [12], and the 2017 revision of the McDonald criteria [13], respectively.

**Table 1:** Participant demographics by type of disease.

| Disease | # Participants | Age (mean ± SD) | Years of Education (mean ± SD) | Gender (ratio) | Paper |
| --- | --- | --- | --- | --- | --- |
| Mild Cognitive Impairment (MCI) | 32 | 71.31 ± 6.83 | 10.54 ± 4.33 | 18 m / 14 f | [14] |
| Multiple Sclerosis (MS) | 18 | 45.05 ± 9.92 | 14.11 ± 4.89 | 6 m / 12 f | [15] |
| Parkinson's Disease (PD) | 20 | 64.5 ± 12.18 | 11 ± 3.9 | 14 m / 6 f | [16] |
| Amyotrophic Lateral Sclerosis (ALS) | 39 | 59.63 ± 12.87 | 10.38 ± 4.3 | 29 m / 10 f | [17] |

Data collection involved MEG and Magnetic Resonance Imaging (MRI) acquisitions. All patients undergo MRI scans on a 3T Biograph mMR tomograph, capturing 3D T1-weighted images. MEG data were acquired using a 163-magnetometer system, with a sampling frequency of 1024 Hz and each patient underwent two 3.5-minute resting-state recordings. Electrocardiographic and electrooculographic signals were also recorded to facilitate artifact removal. Preprocessing steps included Principal Component Analysis (PCA) to reduce environmental noise and Independent Component Analysis (ICA) to remove physiological artifacts. Source reconstruction was performed using a beamformer algorithm on the Automated Anatomical Labeling (AAL) atlas on 116 anatomical Regions of Interest (ROIs). The resulting time series were filtered between 0.5 and 48 Hz. In practice, after preprocessing and artifact removal, we obtained 2-minute recordings of clean signals for each of the 109 patients, sampled at 1024 Hz, across 116 brain regions, yielding a multivariate time series of shape (116, 122880) per subject.

### 2.2 Feature extraction

Given the challenge of identifying reliable and generalizable biomarkers across diverse datasets, we focused on functional connectivity (FC)-based features, which provide a more holistic view of brain function compared to classical local measures where each source is analyzed independently, such as power spectra. Although there are numerous approaches to estimate brain interactions [18], we selected three of the most straightforward methods for simplicity: correlation (Corr), covariance (Cov) and the avalanche transition matrix (ATM).

#### Covariance (Cov)

Covariance matrices provide a robust framework for analyzing multivariate interactions, making them particularly well-suited for the study of brain signals such as EEG and MEG. By capturing the statistical dependencies between multichannel signals [19], covariance matrices enable the application of geometric approaches, a key focus of our work [6]. Formally, a covariance matrix is a symmetric, positive semi-definite *C × C* matrix, where *C* denotes the number of signal channels. Each off-diagonal element (*i, j*) represents the covariance between channel *i* and channel *j*, while the diagonal elements (*i* = *j*) correspond to the variances of the individual channels.

#### Correlation (Corr)

Another way to investigate the statistical relationship between signals consists of computing the correlation matrix. A correlation matrix is a square matrix that expresses the linear correlation between each pair of signals, normalized by their variances. It provides a scale-invariant measure of how strongly signals co-vary. Practically, a correlation matrix is computed from the covariance matrix by normalizing each entry with the product of the standard deviations of the corresponding signal channels. It consists in a it is a semi-definite matrix of rank n-1 (n being the number of considered sources).

#### Avalanche Transition Matrix (ATM)

Another informative feature considered in this study is the ATM, which captures rich, aperiodic, and complementary information relative to conventional measures. Each element (*i, j*) of the ATM encodes the probability that, given region *i* is active at time *t*, region *j* will be active at time *t* + *δ* (typically *δ ≈* 3 ms), as proposed in [20]. ATMs are derived from *neuronal avalanches*, defined as sequences of consecutive time bins during which at least one brain region exceeds a z-score threshold (e.g., *|z| >* 1.6); an avalanche terminates when all regions return to baseline. To construct ATMs, each region’s time series is z-scored and binarized: regions with *|z| >* 1.6 are marked as active (1), and others as inactive (0). For each avalanche, a transition matrix is computed, where element (*i, j*) represents the conditional probability that region *j* becomes active at *t* + 1 given region *i* was active at *t*. Individual avalanche ATMs are then averaged element-wise across all avalanches for each subject and symmetrized, yielding a subject-level matrix that characterizes the propagation of neural activity patterns. Recent insights [14], [16], [17], [21], [22], [23] demonstrated that ATM features may help in the classification by providing a more nuanced representation of the underlying neural dynamics.

### 2.3 Feature dimensionality reduction

The connectivity matrices, of dimension 116 *×* 116, encode pairwise interactions between brain regions, resulting in 6,670 unique features (upper triangle, excluding the diagonal). Given the high dimensionality, feature reduction is essential to mitigate noise, redundancy, and overfitting. For each FC, we employed a refined filtering approach, adapted from [8], to select the most informative connections. This method applies the Kruskal–Wallis test to all pairwise connections, yielding a matrix of effect-sizes. Effect-sizes directly quantify class separability, therefore we used them as the primary criterion for feature selection. A threshold (*effect* − *size >* 0.10) is applied to generate a binary mask. For each brain region (i.e., each node in the FC matrix), we then computed its degree within this binarized matrix. Finally, we retained only the *N_hd_* nodes with the highest degrees. The resulting reduced matrices focus on the most discriminative brain regions, enhancing both classification performance and interpretability. A representation of the filtering process is provided in Figure 1.

**Figure 1:**
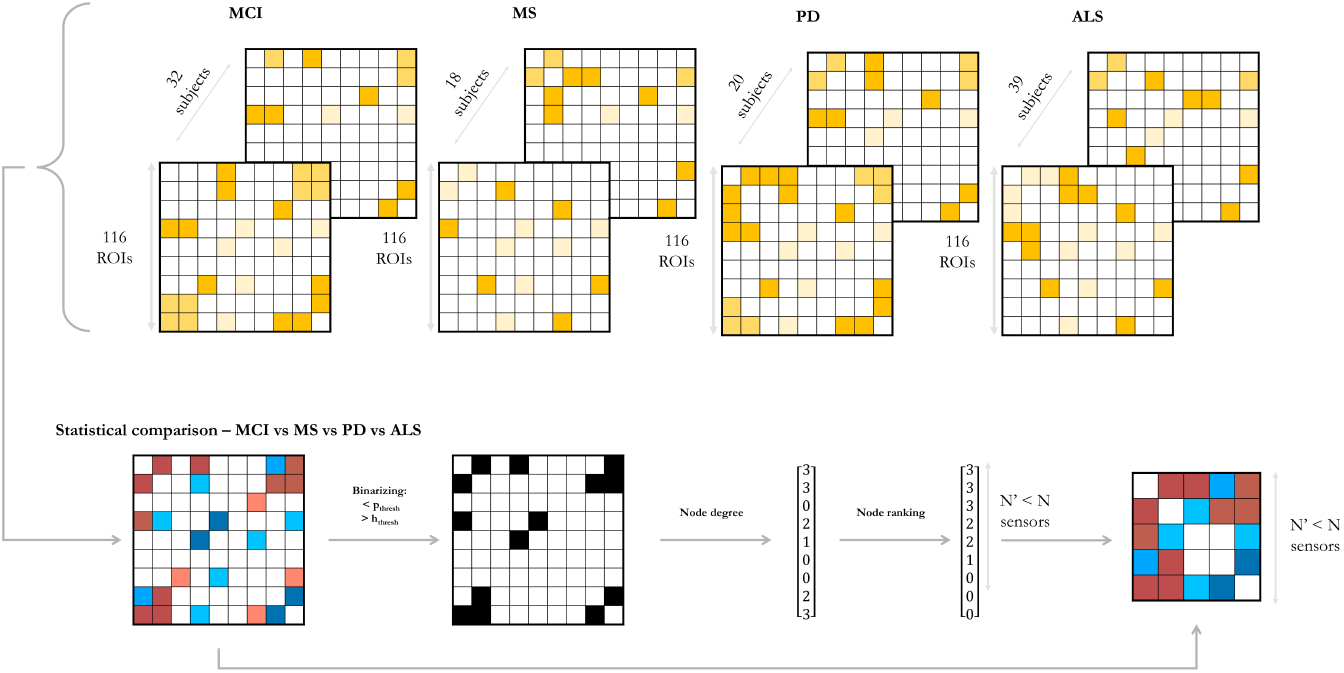
Filtering approach based on the Kruskal–Wallis test. For each connectivity matrix, the test selects the most informative entries.

### 2.4 Modeling and evaluation

In this study, we followed a structured machine learning pipeline guided by two main hypotheses: first, geometry-based methods improve the robustness of the model [6], [24], [25], [26], [27], and second, combining complementary brain-derived features improves classification performance [8].

All the models employed in this study were implemented using TensorFlow [28] (for Neural Networks) and scikit-learn [29], [30] (all other classifiers, including the Riemannian ones), which was also employed for models validation.

#### 2.4.1 Riemannian Geometry Classification

Applying Riemannian geometry to brain signal classification offers several advantages, including high robustness, low noise sensitivity, and minimal calibration requirements [6], [7]. Riemannian classifiers have consistently outperformed Euclidean-based approaches, as evidenced by their success in recent competitions [25], [26], [27]. In this work, we employ the Tangent Space (TS) Classifier, implemented via the pyRiemann library. The core idea is to project symmetric positive definite (SPD) matrices onto a tangent space, enabling the use of standard Euclidean classifiers. Such geometric approaches rely on the assumption that the matrices used to feed the model are SPD. However, numerical inaccuracies may occasionally prevent strict positive definiteness. In such cases, regularization techniques, notably shrinkage methods, are applied to ensure well-conditioned matrices [31], [32].

The space of SPD matrices, denoted by

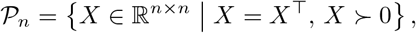

admits a tangent space *T_X_ P_n_* at each point *X* ∈ *P_n_*. The choice of scalar product on the tangent space defines different geometries and associated distances between SPD matrices *X* and *Y*:

- *Euclidean distance:* 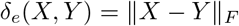
- *Log-Euclidean distance:* 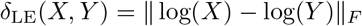
- *Affine-Invariant Riemannian distance:* 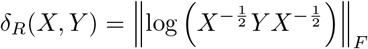

While the Euclidean distance (*δ_e_*(*X, Y*)) treats *P_n_* as a subspace of symmetric matrices, the Log-Euclidean and Affine-Invariant distances are derived from Riemannian geometry, more appropriate for SPD structures.

In our implementation, the covariance, avalanche transition, and correlation matrices were projected into the tangent space, and a *k*-Nearest Neighbors (kNN) classifier, with *k* = 5, was applied in the resulting Euclidean space. This choice was based on preliminary empirical comparisons with other classifiers, as kNN offered the best balance between generalization, performance, and resistance to overfitting.

#### 2.4.2 Ensemble Classification Pipeline

To leverage the complementary information captured by the different estimators, without increasing feature dimensionality, we implemented an ensemble classification framework in which, for each model, two of the available FC matrices are selected and used to independently train two classifiers, each operating on a distinct FC.

We chose to fuse the classifiers’output rather than the different types of features, since the fuse of the output confers higher reliability and robustness through redundancy and facilitates the integration of heterogeneous data without normalizing them [33], [34], [35].

The complete ensemble pipeline is illustrated in Figure 2 and is described below. The extracted features were first independently filtered as described in Section 2.3. The covariance, correlation, and avalanche transition matrices were then projected onto the tangent spaces of Riemannian manifolds, using their class-specific mean as the reference point. In these tangent spaces, two separate kNN classifiers were trained (independently on their respective feature representations; one for each branch of the ensemble). Then, they were used to predict class probabilities for each input data. These probabilities were combined using a confidence-based decision rule: if both models agreed, their common prediction was selected; otherwise, the prediction with the highest confidence (i.e., the maximum class probability) was retained.

**Figure 2:**
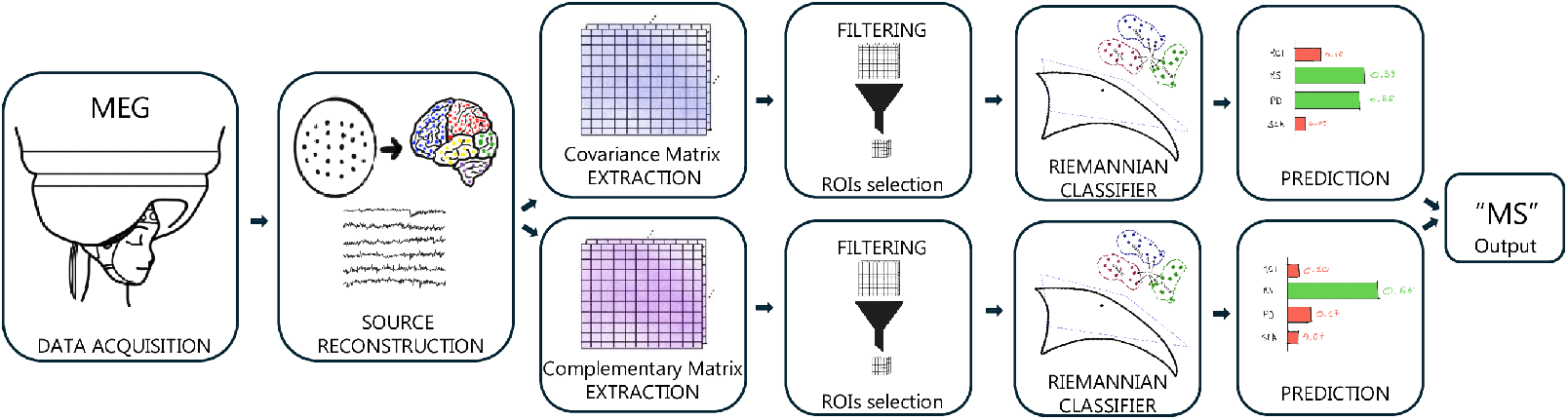
High-level overview of the proposed pipeline. It consists in an ensemble model, in which each classifier is trained independently. The two classifiers employ complementary FC matrices. After applying a filtering approach, a projection in a Tangent Space (TS) is performed, followed by a kNN classification. Predictions are combined using a decision rule, as explained in Section 2.4.2.

Before training the ensembles, the hyperparameters of the kNN classifiers were selected through a preliminary analysis. Specifically, for each functional connectivity matrix type, the corresponding kNN classifier was trained using 5-fold nested cross-validation. For the ATM and Corr matrices, no substantial performance differences were observed across different hyperparameter settings; therefore, the default scikit-learn [29], [30] ones were used (*n*_neighbors_ = 5, metric = ‘euclidean‘, weights = ‘uniform‘). For the Cov matrix, the same configuration consistently achieved the best performance across folds and was thus adopted.

#### 2.4.3 Benchmark

Our approach was compared to other popular features and classification algorithms. To ensure a fair comparison and highlight the efficiency and robustness of Riemannian-based approaches, all the sets of features used to train Riemannian-based classifiers were also used across other classification models, including LDA [36], SVM [37], XGBoost [38] and two Neural Networks [39]. We also considered a wider range of potential features, including Power Spectral Density (PSD) features in the alpha band [40], [41], [42], [43], given the better SNR in these regions. The two neural networks were built as follows: the first network, named Simple NN, consisted of three fully-connected layers [39], trained using the Adam optimizer [44] with a learning rate of 0.001; the second network, referred as Deep NN, consisted of four fully connected layers, with batch normalization layers [45] and dropout layers [46] added after the first two fully-connected ones. The dropout parameter was set to 30% and the Deep NN was trained using Adam with a learning rate of 0.0005.

### 2.5 Validation of analysis

#### 2.5.1 Cross-Validation and Validation Metrics

Given the limited amount of data available in our study (only 109 subjects) and the class imbalance (32 MCI, 39 ALS, 20 PD, 18 MS), each model we used was trained using a 5-fold stratified cross-validation strategy [47], to obtain more reliable performance estimates, enhance robustness and mitigate overfitting. Stratified cross-validation is suited to data-scarce scenarios with class imbalance like ours, as it both allows us to make efficient use of all available data by rotating between training and validation sets while providing multiple estimates of performance variability, while preserving the original proportions of the labels as the original dataset. To avoid data leakage, the feature dimensionality reduction described in sect. 2.3 was performed within the cross-validation loop, using only the training set for feature selection. To capture both the average behavior of the model and its robustness to different data splits, for each metric, we report the mean and standard deviation of performance across folds.

Since our dataset was unbalanced, we avoided relying on plain accuracy, which can be misleading in such contexts [48]. Instead, we evaluated performance using balanced accuracy and F1 score, which are more appropriate to capture both sensitivity and precision across classes. This choice follows best practices for model evaluation in neural engineering, where overestimation of performance is common if care is not taken in metric choice and validation methodology [49].

#### 2.5.2 Statistical Validation

To further validate our analysis and assess statistical differences in performance between pairs of investigated pipelines, we employed statistical testing. All statistical analyzes presented in the subsequent sections used a one-tailed Mann–Whitney U test [50], a nonparametric test suitable for small sample sizes. In our case, each population consists of five samples, corresponding to the five cross-validation folds. We implemented statistical tests via the scipy library [51].

## 3. Results

### 3.1 Selected features

Before starting the differential classification analyses, we first inspected the global output of the filtering procedure. Using the full dataset, we computed the rank of each brain region, defined as its position in the list of nodes sorted in descending order by the supra-threshold effect sizes computed on the pairwise connections (please refer to 2.3 for the filtering procedure). In Figure S1, each region is color-coded according to its rank. To illustrate the behavior of the filtering procedure during cross-validation, we report in Figure 3 the mean rank of each ROI, averaged across the 5 folds, while Figure S2 how often a given node was selected across folds. All of these visualizations are provided for the covariance, correlation, and avalanche transition matrices (ATM). Comparison of Figure 3 with its counterpart generated using the complete dataset (Figure S1) reveals a very high degree of concordance, demonstrating that the filtering procedure is highly robust to the choice of folds. The most informative regions, for the correlation matrix, were the right insula, the right Heschl’s gyrus and the left precuneus, together with bilateral inferior parietal cortices, supplementary motor areas, paracentral lobules, postcentral cortex, and mid-cingulate regions. For the covariance matrix, the right insula again emerged as the most discriminative node, followed by the left supplementary motor area, the right parahippocampal gyrus, and the right paracentral lobule. The remaining highly ranked regions included bilateral temporal poles, lingual and occipital cortices, fusiform gyrus, Rolandic operculum, cingulate cortices, inferior frontal regions, precuneus, and precentral/postcentral areas. For the ATM-based connectivity, the most informative regions included the right inferior parietal cortex, left mid-cingulate cortex, right supramarginal gyrus, left insula, right calcarine cortex, bilateral cuneus, medial superior frontal cortex, bilateral postcentral cortex, Rolandic operculum, precentral cortex, and supplementary motor area. Across all three connectivity definitions, a consistent pattern emerges: sensorimotor (precentral, postcentral, paracentral lobule, supplementary motor area), inferior parietal/supramarginal, insular, and mid-cingulate regions repeatedly rank among the most discriminative nodes. Occipital regions (calcarine, cuneus, lingual, inferior occipital) also appear prominently, particularly for covariance and ATM measures. Importantly, motor impairment represents a predominant clinical feature in three of the four disorders considered (PD, ALS, MS), lending neurobiological plausibility to the prominence of motor and peri-motor cortices. The additional involvement of insular, cingulate, and parietal associative regions further suggests that disease discrimination relies on distributed reorganization of the network rather than on isolated focal alterations.

**Figure 3:**
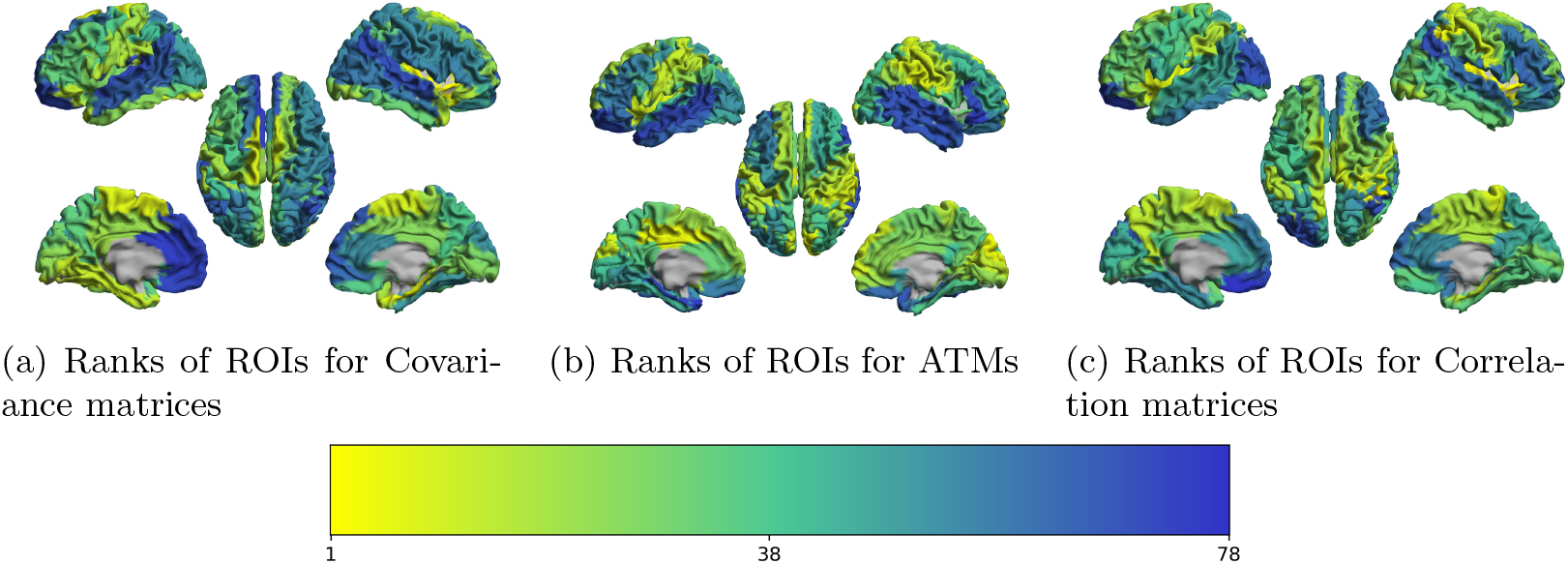
Brain plots showing the mean ranks averaged among the five folds for each ROI. ROIs with a lower average rank are shown in yellow, while the ROIs with higher rank are shown in blue.

### 3.2 Effect of estimators

The first step involved identifying the FC estimators and the corresponding optimal number of nodes *N_hd_* that led to the best performances. We considered the estimators defined in section 2.2 and compared the accuracy obtained with the riemannian approach, while varying the number of retained nodes *N_hd_∈ {*20, 35, 50*}*.

For each of the considered matrices, the analysis revealed that optimal performance was achieved for a value of selected nodes *N_hd_* = 50 (see Supplementary Table S1). Therefore, when not explicitly mentioned, we will refer to this value in all subsequent analyses. The corresponding performance values are reported in Figure 4.

**Figure 4:**
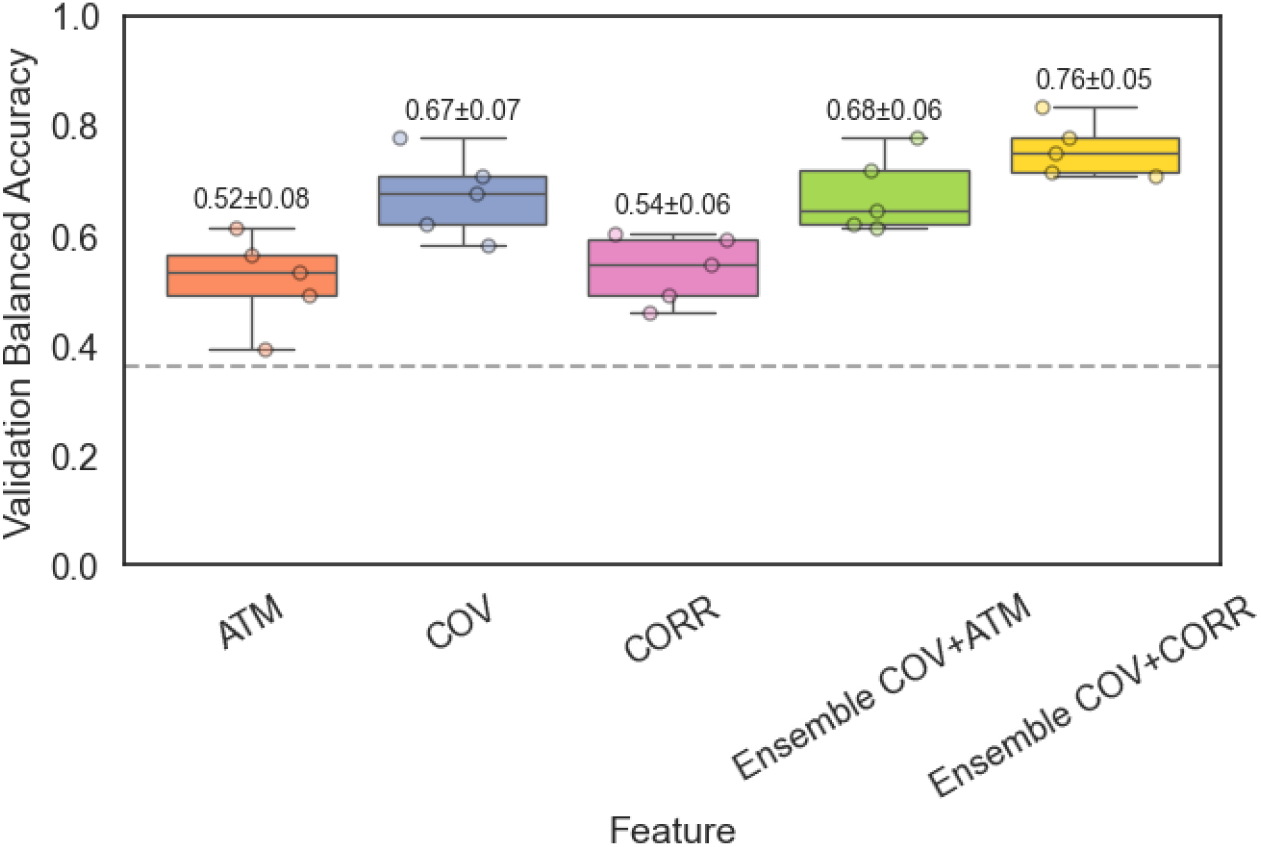
Validation Balanced Accuracies of the Riemann (TS+KNN) method, with *N_hd_* set to 50. Individual scatter points indicate the performance on each fold, while boxplots summarize the distribution across folds, including values corresponding to extremes, quartiles, and the median. The Cov+Corr configuration achieves the best performance. The dashed line indicates the chance level.

Covariance was the best-performing estimator, achieving a balanced accuracy of 0.67 *±* 0.07. Specifically, Cov significantly outperformed both Corr (0.54 *±* 0.06; *p* = 0.016) and ATM (0.52 *±* 0.08; *p* = 0.008).

We also investigated whether combining single-level classifiers could improve performance. Since covariance was the best-performing FC estimator when used alone, it was always included when constructing the ensembles.

The ensemble configurations showed balanced accuracies of 0.76 *±* 0.05 and 0.68 *±* 0.06, corresponding to the combinations Cov+Corr and Cov+ATM, respectively (see Figure 4). Among all the configurations explored, the ensemble combining the covariance and correlation estimators was the best-performing. This configuration outperformed not only the stand-alone estimators (being significantly better than ATM, Cov, and Corr, with *p*-values *<* 0.005), but also the alternative ensemble classifier Cov+ATM, for all *N_hd_* explored.

As a result, we worked with the configuration presented in Figure 4 that includes the estimation of the covariance, and the correlation. We also note that this configuration, combined with selecting *N_hd_*= 50 nodes, achieved the best performance among all the considered scenarios (that is, varying features and *N_hd_*). This includes the case without node selection, where all 78 nodes were retained (balanced accuracy of Cov+Corr 0.71 *±* 0.10). In the following sections, we use pipeline REDDI, to indicate the configuration shown in Figure 4 (corresponding to the ensemble approach relying on the Cov+Corr).

### 3.3 REDDI, a robust and interpretable approach for differential diagnosis

We compared the performance of our REDDI pipeline against other popular classifiers (see Figure 5). As previously mentioned, the same set of features was used across all classification models to benchmark them, and *N_hd_*= 50 has been selected (for performance comparisons with the other cases, please refer to Supplementary Figure S3). The REDDI pipeline achieves the highest balanced accuracy (0.76 *±* 0.05). We list the balanced accuracies of the other pipelines in Figure 5, and report in table 2 the p-values resulting from the test of whether REDDI significantly outperforms them in terms of balanced accuracy.

**Figure 5:**
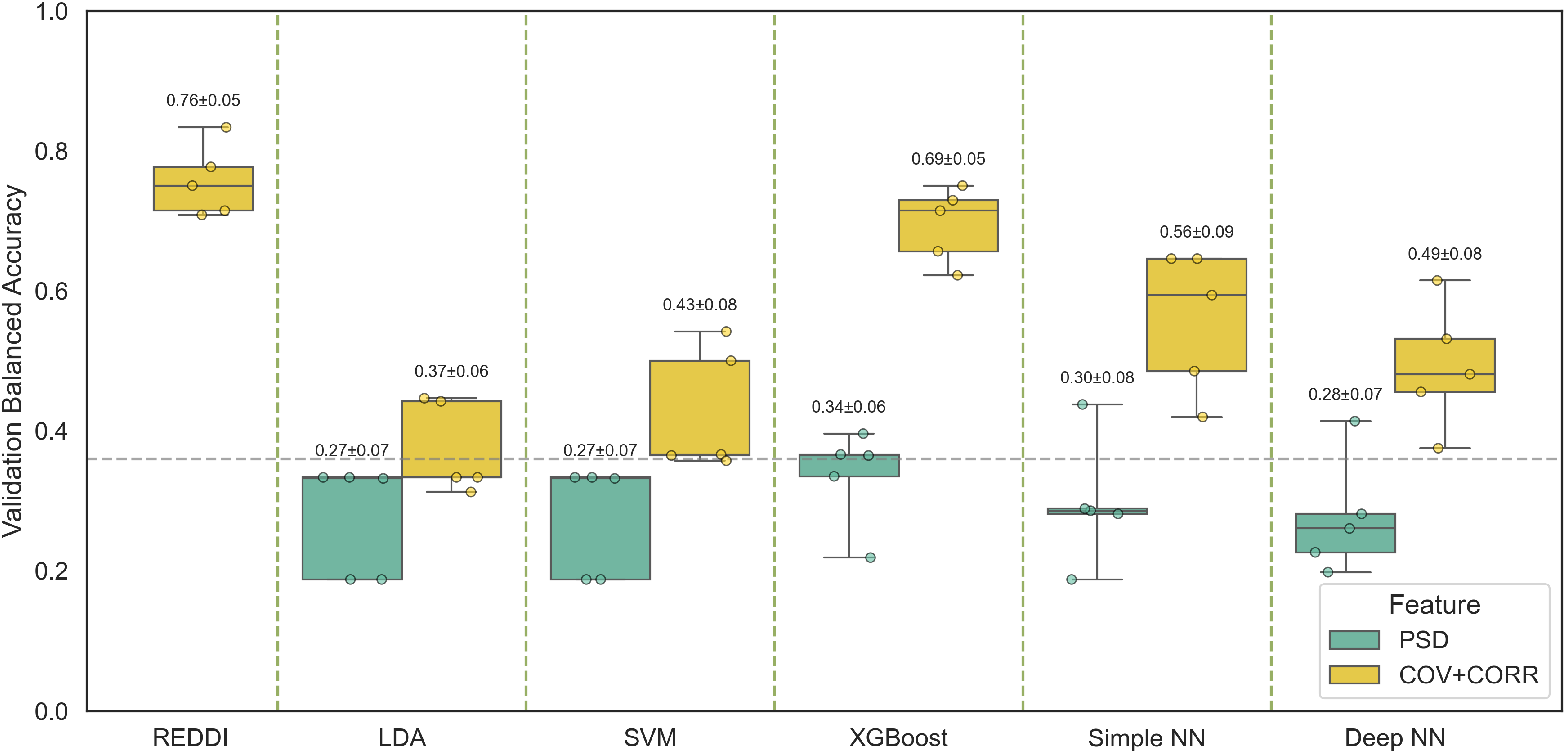
Benchmark: Balanced Accuracies of our proposed method (REDDI) compared to other popular classifiers. Individual scatter points indicate the performance on each fold, while box-plots summarize the distribution across folds. REDDI (our method) achieves the best performance. The dashed line indicates the chance level.

**Table 2:** Performance of different pipelines compared to REDDI (0.76 *±* 0.05). The p-values indicate statistical difference from REDDI.

| REDDI vs PSD |  | REDDI vs Cov+Corr |  |
| --- | --- | --- | --- |
| Pipeline | p-value | Pipeline | p-value |
| LDA | 0.006 | LDA | 0.006 |
| SVM | 0.006 | SVM | 0.004 |
| XGBoost | 0.004 | XGBoost | 0.1 |
| Simple NN | 0.004 | Simple NN | 0.006 |
| Deep NN | 0.004 | Deep NN | 0.004 |

Figure 5 clearly shows that REDDI outperforms all other pipelines. Table 2 shows that the performance of REDDI is significantly better (*p <* 0.01) than that of any other pipeline, except for XGBoost (Cov+Corr case). Although the improvement over XGBoost is less statistically significant, as shown in Supplementary Fig. S4 the Riemannian models are the only ones that do not exhibit signs of overfitting when comparing training and validation performance, highlighting the advantage of our REDDI pipeline over XGBoost. By contrast, other models, particularly LDA and XGBoost, show a substantial performance gap, a clear indication of overfitting. This demonstrates the generalization advantage of the Riemannian’s approaches and, in particular, of REDDI.

A more comprehensive benchmark is provided in Supplementary Figures S4 and S5, which show the balanced accuracies and F1 macro scores, respectively, for the combination of the six classifiers (LDA, SVM, XGBoost, Riemannian (TS+kNN), Simple NN, and Deep NN) and the six features (PSD, ATM, Cov, Corr, Cov+ATM, Cov+Corr). Among all model and feature combinations, the ensemble of covariance and correlation matrices (Cov+Corr) within the Riemannian framework achieves the highest performance with no sign of overfitting, demonstrating that REDDI is the overall best pipeline.

Given the superior performance of REDDI, we further explored its explainability. To this end, we built confusion matrices (6a) to visually inspect the distribution of errors across classes. We also reported the average prediction probability belonging to the predicted class for each predicted–true class combination in the confusion matrix to estimate the confidence of correct and incorrect classifications (6b). Finally, we provided ROC curves in (6c). The confusion matrix are derived from out-of-fold predictions during cross-validation. The model demonstrates strong performance across most classes, except for PD, which exhibits a higher misclassifications rate. Complementing the confusion matrix, the average probability matrix illustrates the model’s mean confidence for each true– predicted class pair. From this matrix, we calculated the hit confidence—the average predicted probability for correctly classified samples (i.e., the diagonal of the probability matrix), which was 82% (*±*14). In contrast, the missed confidence, i.e., the average probability assigned to incorrect lowered to 63% (*±*9). Finally, the ROC curves (shown in Figure 6c) reveal near-perfect classification for MS, with an AUC of 0.99, while PD shows the weakest performance. The step-like shape of the ROC curves arises because the model outputs only five distinct probability values, resulting in a limited number of thresholds.

**Figure 6:**
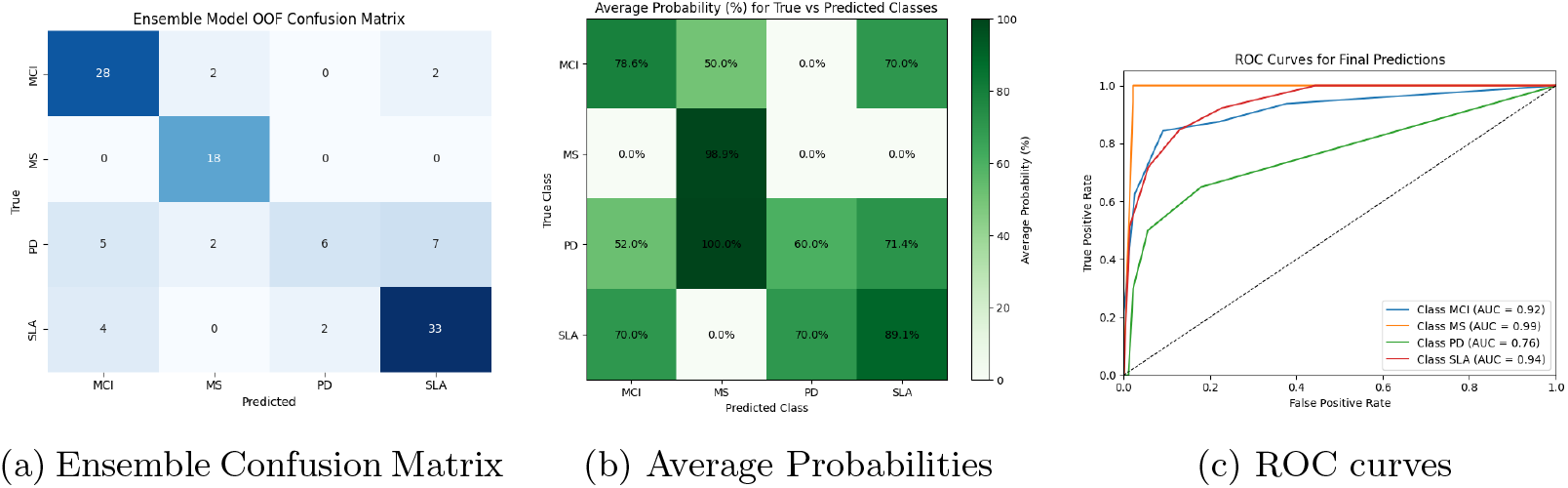
Confusion matrix (a), average probabilities (b), and ROC curves (c) for REDDI.

## 4. Discussion

In this study, we proposed and evaluated the REDDI approach, a classification pipeline for differential diagnosis (among MS, MCI, PD, and ALS) that leverages Riemannian geometry-based classifiers and novel features such as Covariance Matrices and weighted Avalanche Transition Matrices (ATMs). To maintain interpretability while reducing feature dimensionality, we implemented a model-agnostic feature selection process using Kruskal-Wallis statistics and effect-size filtering. This approach successfully reduced the number of considered features by 81%.

To assess the potential of REDDI for differential diagnosis, we conducted a comprehensive benchmark analysis considering alternative features (specifically PSD) and classification tools (including SVM, LDA, XGBoost, and both simple and deep neural networks). The primary objective of our research was to develop interpretable neurophysiological tools for differential diagnosis using machine learning techniques, with a particular focus on Riemannian classifiers applied to source-reconstructed MEG data. Our approach utilized features such as covariance, correlation, and avalanche transition matrices to preserve interpretability while enhancing diagnostic effectiveness. The REDDI pipeline achieved an accuracy of 0.76.

### 4.1 Interpretation of discriminating features

Across the three metrics, a remarkably consistent pattern emerges: the most discriminative regions are not confined to a single canonical network but cluster at anatomically central and metabolically demanding nodes, highly connected and therefore potentially more vulnerable to network-level perturbations, regardless of the disease-specific noxae. Convergence on the bilateral insula, mid-cingulate cortex, supplementary motor area (SMA), paracentral lobule, inferior parietal cortex, precuneus, and para-hippocampal regions suggests that multi-disease classification is driven less by focal, disease-specific epicentres and more by hubs that mediate large-scale integration. Functionally, these regions have often been related to higher-level functions, such as, for example, interoception and salience detection (insula), motor planning and initiation (SMA, paracentral lobule, pre/postcentral gyri), and sensory integration (inferior parietal, supramarginal). On the one hand, the prominence of such regions might support the idea that they might act as “integration bottlenecks”, where distinct molecular pathologies converge onto shared systems-level dysfunction. On the other hand, these are regions that sit at the interface between motor, cognitive, and limbic domains, which are some of the dimensions that differentially combine across the disorders taken into account. In other words, these regions are not merely disease markers; they are multiscale control nodes whose differential impairment patterns can encode the signature of multiple conditions. This interpretation is consistent with the findings reported in [5], in which phase-based edge metrics outperformed nodal measures in multi-disease classification. While the techniques utilized in the two works differ and do not allow a straightforward comparison, the edges identified as most informative hinge on nodes that are remarkably similar to those in the current work. The previously identified superiority of the edge metrics and the importance of integrative territories highlighted in the current work suggest the idea that distinct neurodegenerative processes might reshape large-scale dynamical interactions, most prominently in highly interconnected regions.

### 4.2 Comparative Analysis and Performance

Our findings align with and extend previous research [5], [52], [53], [54], [55], [56], indicating that brain signals, particularly MEG recordings, contain discriminative information crucial for recognizing, classifying, and categorizing neurodegenerative diseases. This suggests that these conditions exhibit distinct biomarkers that affect brain function at a global scale [57], [58], [59]. Within the existing literature, some studies have focused on the biological and molecular mechanisms underlying neurodegenerative diseases, such as the work by [60]. Others have aimed at differential diagnosis similar to our project but used structural MRI or EEG data to distinguish between various neurodegenerative conditions, subtypes, and variations [61], [62]. These results collectively highlight the effectiveness of machine learning in classifying neurological disorders, supporting their broader application in clinical diagnostics (see Table 3). However, our study advances this approach by addressing the challenge of multi-disease classification, which is more representative of real-world clinical scenarios.

**Table 3:** Comparison of classification performance in existing literature.

| Data (n) | Disease(s) | Features | Method | Results | Source |
| --- | --- | --- | --- | --- | --- |
| MEG<br>(n=233) | 67 Control,<br>26 SCI, 140<br>Epilepsy | MEG time se-<br>ries | MNet (DNN)<br>and SVM | Accuracy:<br>70.7% ± 10.6<br>(vs. 63.4%<br>± 12.7 with<br>SVM); Speci-<br>ficity: 86-94% | [63] |
| EEG<br>(n=295) | 180 AD, 115<br>MCI | From temporal<br>data to com-<br>pressed images | MLP and AHR<br>networks | Accuracy:<br>92.33% (AHR);<br>90.82% (MLP) | [64] |
| EEG<br>(n=141) | 52 Control, 52<br>AD, 37 MCI | Topographical<br>images, time-<br>frequency<br>graphs | AlexNet (DL)<br>with 10-fold<br>CV | Accuracy:<br>98.9% ± 0.4 | [65] |
| EEG<br>(n=242) | 45 Control, 116<br>Mild AD, 81<br>AD | Time, fre-<br>quency, com-<br>plexity, PSD,<br>connectivity | Random For-<br>est, SVM,<br>FFNN | ML: Sen-<br>sitivity:<br>85%, Speci-<br>ficity: 78%;<br>SVM/NN:<br>Sensitivity:<br>89%, Speci-<br>ficity: 88% | [66] |
| EEG<br>(n=48) | 15 Control, 17<br>Early AD, 16<br>Early MCI | rPSD and com-<br>plexity features | Binary SVMs<br>with Leave-One-Out CV | Accuracy:<br>83.3%, 85.4%,<br>and 79.2% | [67] |
| rs-fMRI<br>(n=30) | 10 Control, 10<br>AD, 10 MCI | Coh, iCoh,<br>PLV, SL, PAC,<br>AEC | ANOVA statis-<br>tical analysis | Significant<br>group differ-<br>ence, e.g., $p =$<br>0.001 for Beta<br>P7-P8 | [68] |
| EEG<br>(n=295) | 120 Control,<br>175 AD | Small-World<br>values by<br>rhythm | PCA + SVM<br>with repeated<br>CV | Accuracy: 95%<br>± 3; Sensitiv-<br>ity: 95% ±<br>5; Specificity:<br>96% ± 3; AUC:<br>0.97 ± 0.03 | [69] |

### 4.3 Advantages of the REDDI Approach

The effectiveness of our REDDI architecture is largely attributable to the robustness of its individual components. The flexible, model-agnostic filtering method adapted from [8] ensures adaptability across varying experimental contexts, while the use of effect sizes as a feature selection criterion is particularly well-suited to classification tasks, as it directly quantifies the magnitude of distributional shifts between conditions, thereby prioritizing the most statistically relevant connectivity features.

The superior classification performance observed with covariance and correlation matrices over the ATM may be attributed to several complementary factors. First, covariance and correlation capture distinct yet synergistic aspects of neural dynamics. Covariance preserves both regional signal variance and inter-regional co-activation, thereby reflecting changes in neural power alongside functional coupling. Correlation, by normalizing for signal amplitude, isolates the topological structure of functional connectivity independently of power fluctuations, providing the classifier with sensitivity to pure network reorganization. Second, unlike covariance and correlation, which retain the full continuous structure of the signal, the ATM requires binarization of the data based on a predefined threshold, a step that introduces an inherent loss of information. This discretization may discard subtle yet diagnostically relevant graded fluctuations in neural activity. We therefore hypothesize that, for the pathological conditions investigated here, the most discriminative features are encoded in the continuous dynamics of functional connectivity rather than in the discrete, avalanche-based propagation patterns that the ATM is specifically designed to characterize.

Beyond the choice of connectivity metrics, the Riemannian framework itself conferred important advantages over conventional machine learning approaches, including XGBoost, Gaussian Naive Bayes (GNB), Linear Discriminant Analysis (LDA), Support Vector Machines (SVM), as well as shallow and deep neural networks, all of which exhibited signs of overfitting. In contrast, our approach achieved superior classification performance while maintaining stability across cross-validation folds, as evidenced by low standard deviations and consistent training and validation performances.

A key strength of the proposed ensemble strategy is that it enables the integration of complementary information sources without inflating the dimensionality of the problem. This is particularly relevant in the context of Riemannian geometry, where performance is known to degrade as feature dimensionality increases. Indeed, it has been shown [6] that as the number of sensors grows, and consequently the size of the covariance matrices, classification accuracy declines, likely due to the increasing number of samples required to estimate non-singular covariance matrices in high-dimensional spaces. Our ensemble approach circumvents this limitation by performing parallel computations on separate, independently filtered feature sets, each operating at a fixed and manageable dimensionality. The predictions derived from these parallel classifiers are subsequently combined for the final decision, thereby leveraging the complementary information encoded in different connectivity representations without compromising the geometric assumptions underlying the Riemannian framework.

### 4.4 Intepretability

Beyond performance improvement, investigating and challenging the interpretability of REDDI is crucial for ensuring its clinical relevance. The model demonstrates strong performance across most classes, except for Parkinson’s disease (PD), which exhibits a higher misclassification rate. The ROC curves confirmed these trends, showing near-perfect classification for Multiple Sclerosis (MS) with an AUC of 1.0, while PD showed the weakest performance.

This disparity might be due, in part, to the fact that Parkinson’s disease originates primarily in deep brain regions, which contribute significantly less to the MEG signals are generate less reliable signals. Consequently, we excluded deep sources from feature selection due to their poor signal quality. Complementing the confusion matrix, we investigated the average probability matrix. The hit confidence was 82%, while the miss confidence was 63%. This difference suggests that the model is generally more confident in its correct predictions than in its errors, which is a positive indicator of model robustness.

### 4.5 Comparison with our previous contribution

We successfully enhanced the performance of the existing machine learning pipeline using Riemannian classifiers. The improvement in our results can be attributed to two main factors.

First, we robustified the feature filtering approach compared to our previous work [5]. Rather than using the presence of a potential statistical difference between distributions via the p-value, we considered the normalized magnitude of distribution shifts—that is, the effect-size—which allowed us to reduce dimensionality while preserving interpretability. This approach is more directly aligned with the goal of maximizing class separability in a classification context. Unlike the approach taken in [5], we did not work with phase-based connectivity estimators, as our aim was to propose a classification tool that could be interpretable in broad-band to capture richer information from brain dynamics.

Second, these results confirm the efficiency and capability of Riemannian classifiers in extracting and fusing relevant information from Symmetric Positive Definite (SPD) matrices [24]. The balanced accuracy of our approach (ensemble of Riemannian classifiers) out-performs other approaches, notably the method we proposed previously [5], which shared the same dataset and preprocessing methods. While the earlier study achieved a best balanced accuracy of 0.67, our method reached a balanced accuracy of 0.76, representing a 9% improvement over the state of the art.

### 4.6 Limitations & Future Perspectives

This study presents several limitations that must be acknowledged. First, the dataset used is relatively small, consisting of only 109 subjects. Our methodology required the implementation of statistical techniques to address the inherent challenge of class imbalance, as reflected by the uneven distribution of sample sizes across diagnostic groups (*N*_ALS_ = 39, *N*_MCI_ = 32, *N*_PD_ = 20, *N*_MS_ = 18). While the observed sample distribution corresponds to real-world prevalence rates, it is critical to recognize that such imbalance may introduce systematic bias into the classification process, potentially compromising the robustness and generalizability of the findings.

This limited and uneven sample size poses significant challenges for both training and evaluation. As an independent test set was not available, all evaluations were based on cross-validation or its variations (e.g., repeated CV). While care was taken to ensure that validation data remained strictly separate from training data in each fold, the absence of a completely unseen external dataset may limit the generalization of our findings. To address these challenges, alternative strategies for managing imbalanced datasets have been explored in the literature. Undersampling the majority class, although effective in balancing class representation, may inadvertently reduce the overall sample size, thereby increasing the signal-to-noise ratio. Conversely, oversampling the minority class through data augmentation techniques [70], [71], [72] can enhance the representation of underrepresented classes but may also introduce noise and elevate the risk of overfitting [73].

A particularly promising avenue to address class imbalance involves leveraging generative artificial intelligence to synthesize data and produce more balanced training sets. This approach, investigated in recent studies [74], [75], has the potential to improve the reliability of classification models by generating synthetic observations that preserve the statistical characteristics of the original data, thereby enhancing the robustness of the classification framework without requiring additional data collection.

Furthermore, while Riemannian geometry offers numerous practical advantages, including robustness and low sensitivity to noise, interpreting the decisions made by Riemannian classifiers remains challenging. Current efforts are therefore focused on developing interpretable frameworks tailored to this geometric setting, aiming to provide deeper insight into the features driving classification [76], [77]. Finally, we are currently extending our benchmarking to include deep learning approaches [78], [79]. Although such architectures are often considered black-box models, where decisions are difficult to interpret and trace back to neurophysiologically meaningful features, the model-agnostic nature of our filtering pipeline ensures that the input features fed into these models remain both statistically relevant and neurologically interpretable.

## 5. Conclusion

In this study, we presented and validated an interpretable machine learning pipeline for the differential diagnosis of four neurodegenerative diseases, mild cognitive impairment (MCI), multiple sclerosis (MS), Parkinson’s disease (PD), and amyotrophic lateral sclerosis (ALS), using resting-state magnetoencephalography (MEG) data. Our central hypothesis posits that integrating geometry-based models with covariance and correlation matrix features would improve diagnostic performance while maintaining interpretability.

Leveraging Riemannian classifiers, we demonstrated that tangent space projections significantly enhanced classification outcomes. Feature selection was further refined using the Kruskal–Wallis test and the effect-sizes. The proposed ensemble model combined predictions from both covariance- and correlation-based classifiers, achieving a mean balanced accuracy of 0.76 *±* 0.05 via 5-fold cross-validation, a notable improvement over the previous benchmark of 0.67. The model also exhibited robustness and low overfitting, as evidenced by the small standard deviation across folds.

These findings underscore the potential of our framework as a robust, interpretable tool for MEG-based diagnosis, with promising implications for future clinical decision-support systems.

## Data Availability

MEG recordings used in this study are not publicly deposited, as open sharing was not covered under the original ethical approval.

## 6. Code availability

The code used to perform the analysis and the data that support the findings of this study are publicly available in this Github repository: https://github.com/mccorsi/REDDI.

## Acknowledgments

This research was supported by the Italian National Recovery and Resilience Plan (PNRR) —European Union —Next Generation EU, DM 118/2023, CUP D42B23001820006 which funded the PhD fellowship of Giovanni Messuti.

## Supplementary

**Figure S1:**
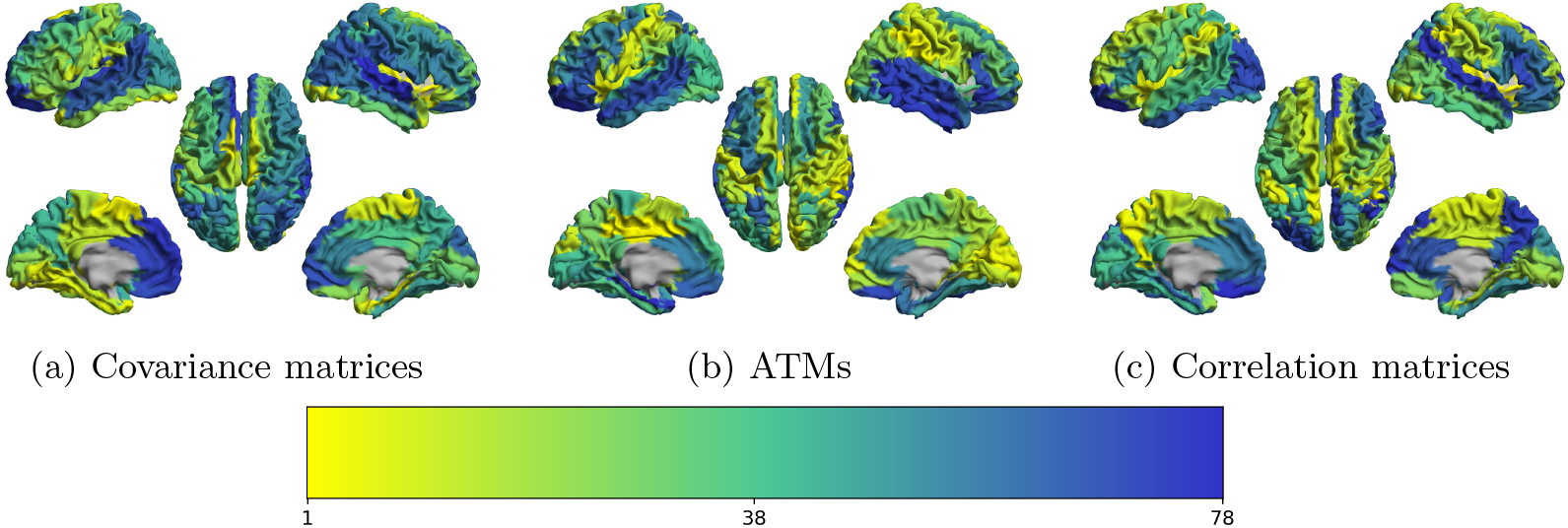
Brain plots showing the ranks of each ROI considering the whole dataset. Each region is colored according to its rank. Further details on the computation of ranks are provided in Section 2.3

**Figure S2:**
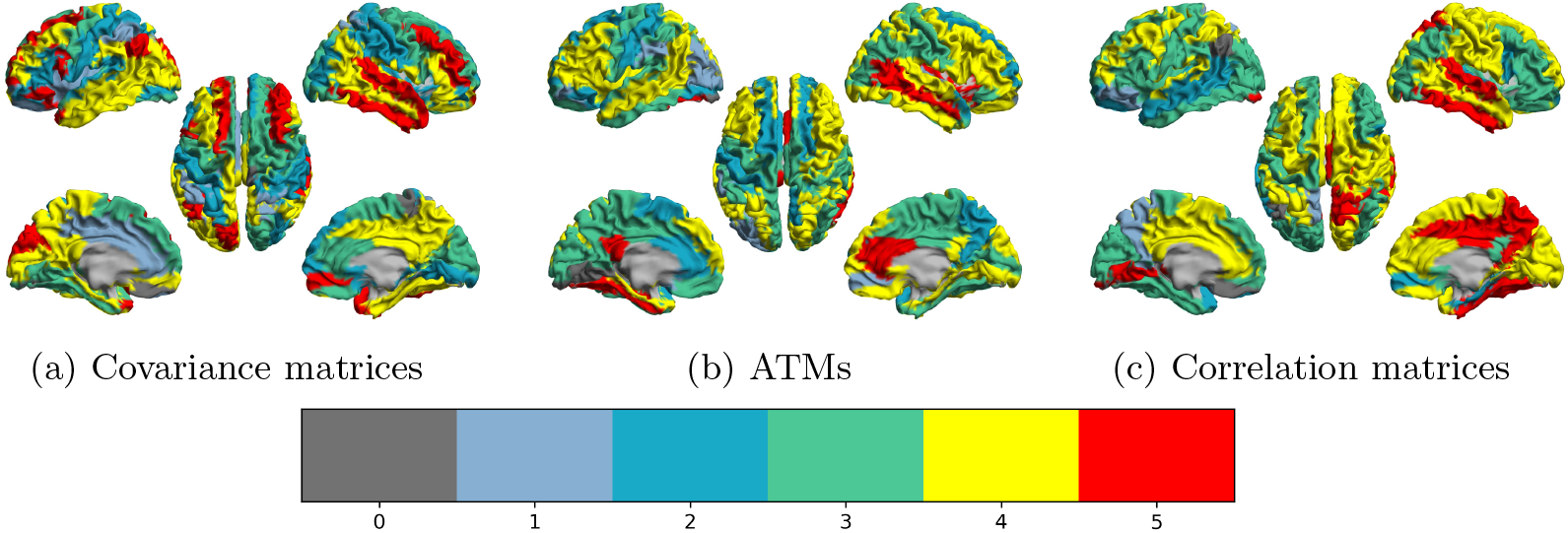
Brain plots showing how many times a region has been selected among the 5 folds by the filtering approach. We recall that a ROI is selected if its rank is lower than 50.

**Figure S3:**
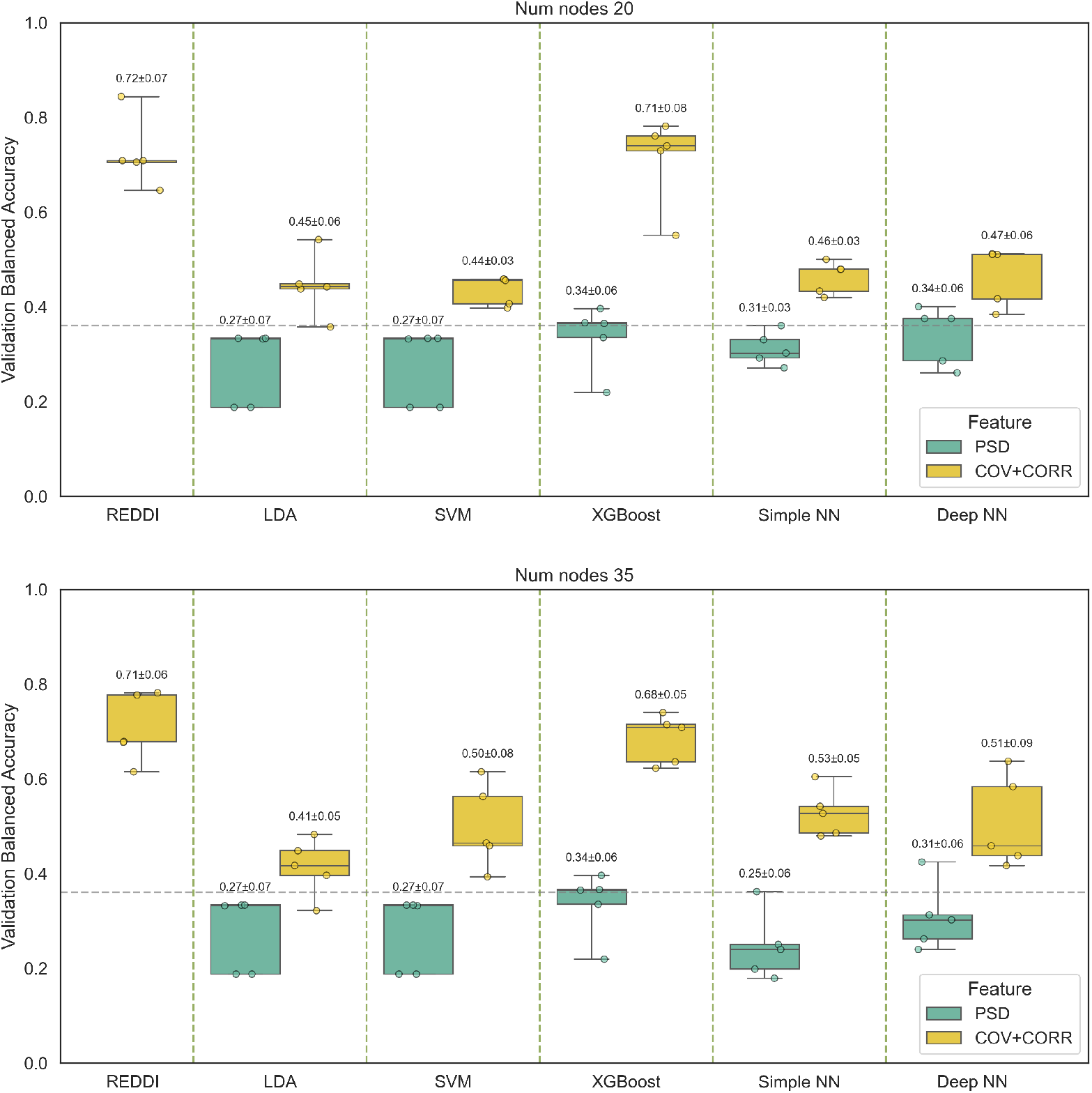
Balanced Accuracies of our proposed method REDDI compared to other popular classifiers, while varying the number *N_hd_* of selected nodes (*N_hd_*=20 on the top and *N_hd_*=35 on the bottom). Individual scatter points indicate the performance on each fold, while boxplots summarize the distribution across folds. The dashed line indicates the chance level. For each *N_hd_* REDDI method achieves the best performance. We recall that the highest overall performance (0.76 *±* 0.05) was achieved with the REDDI approach when selecting *N_hd_*=50.

**Figure S4:**
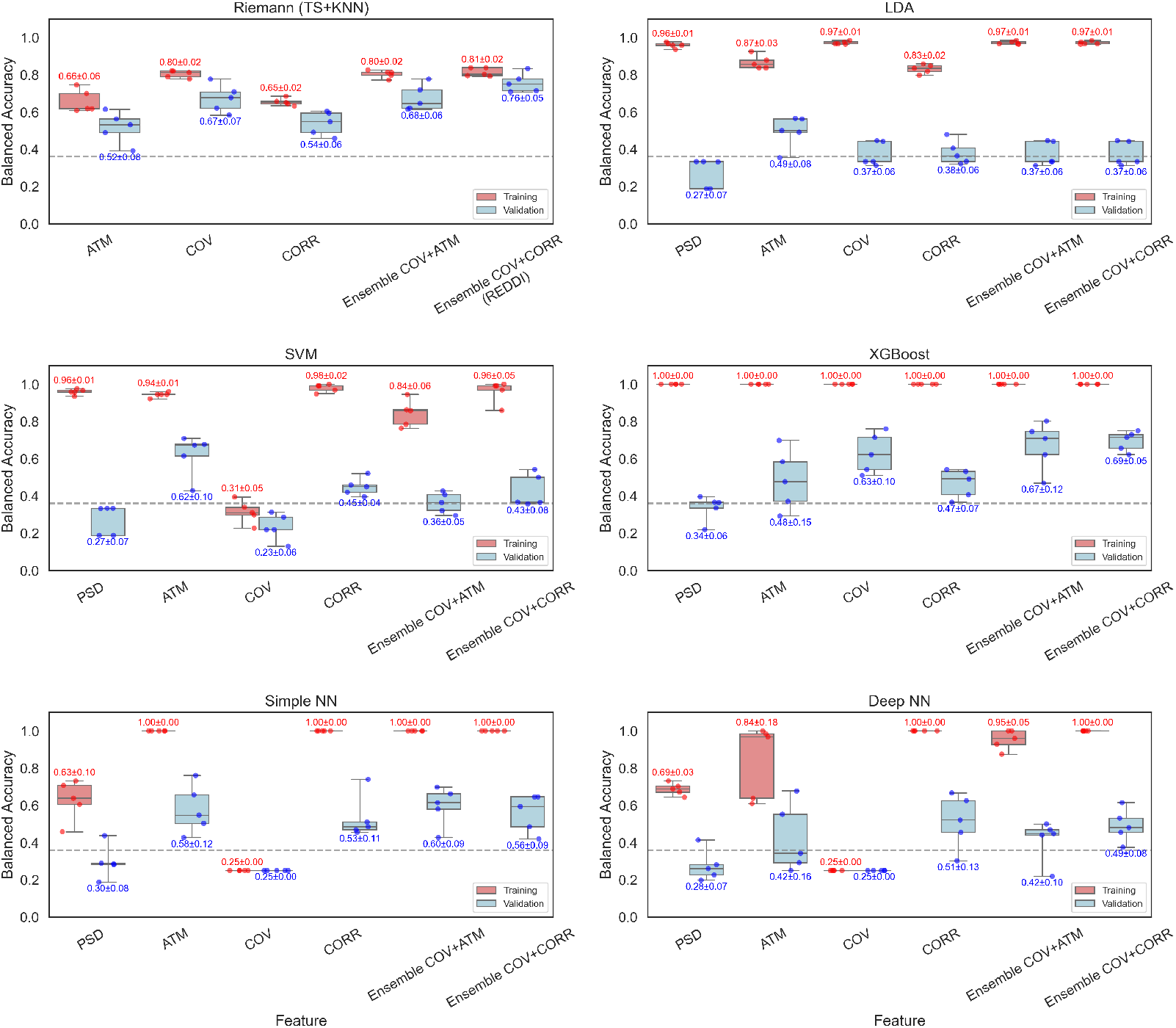
Performance of extended benchmark, comparing REDDI (corresponding to the setting Reimann - Ensemble COV+CORR) with other approaches. Training and Validation Balanced Accuracy are shown. We show *N_hd_* = 50

**Figure S5:**
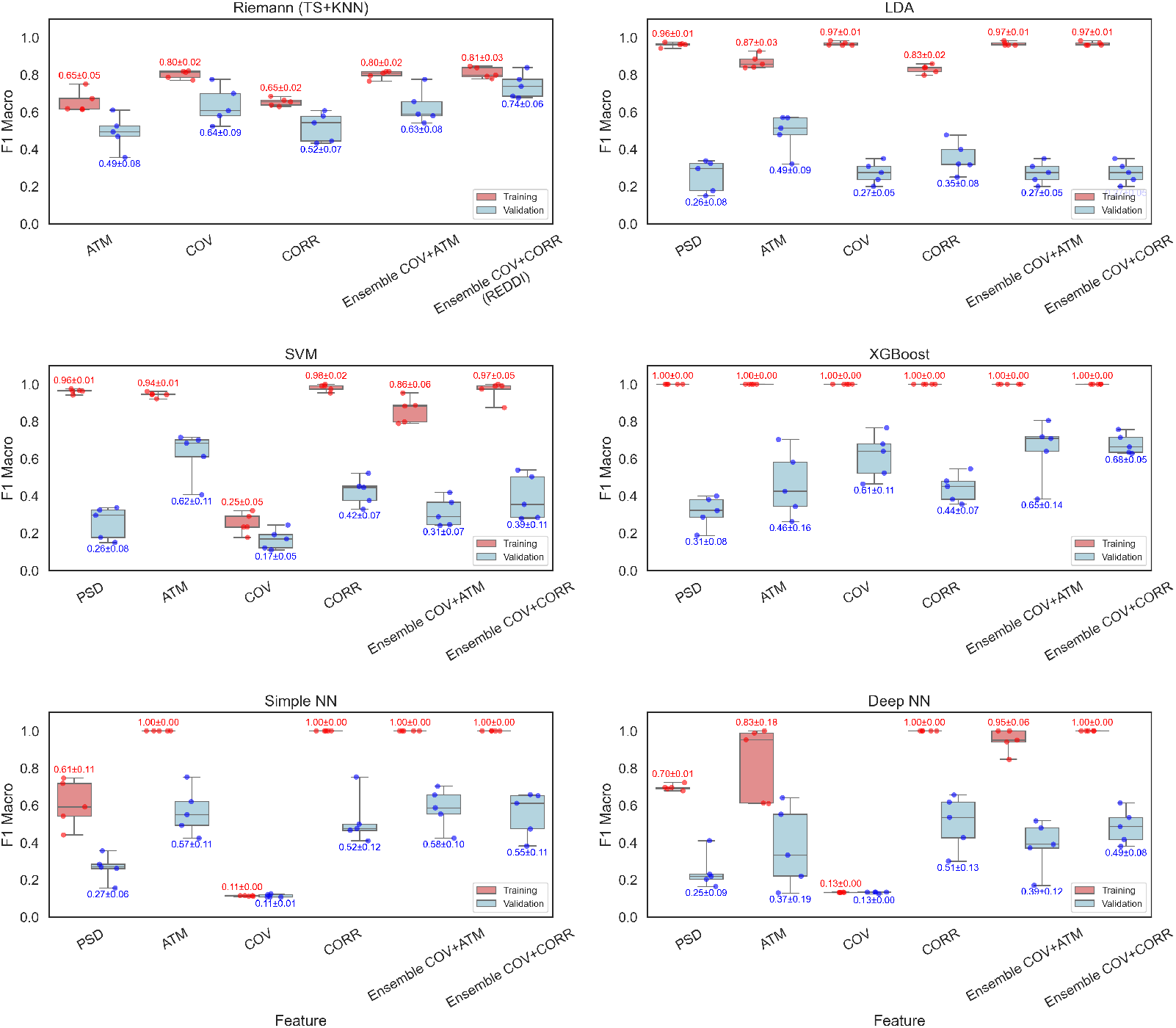
Performance of extended benchmark, comparing REDDI (corresponding to the setting Reimann - Ensemble COV+CORR) with other approaches. Training and Validation F1 scores are shown. We show *N_hd_* = 50

**Table S1:**
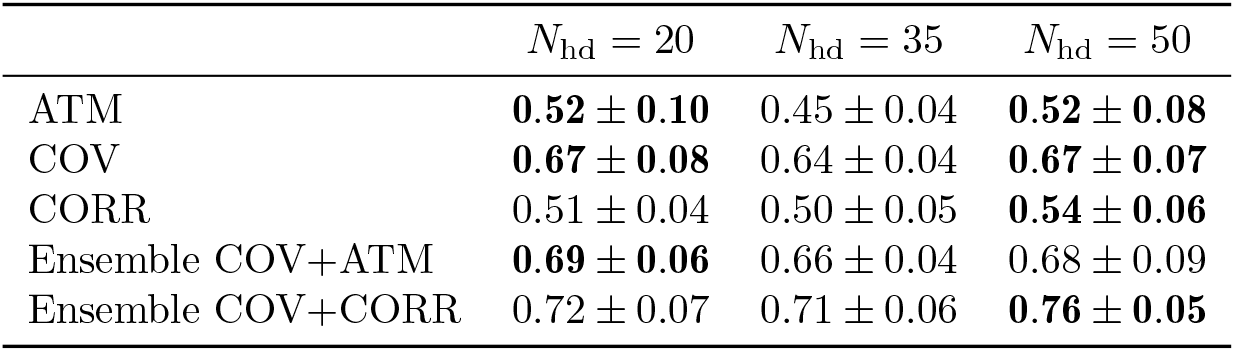
Validation balanced accuracies of the Riemann (TS+KNN) method, varying *N*_hd_. Values are reported as mean *±* standard deviation. For each (single) FC, the best performance, shown in bold, is achieved when *N*_hd_
= 50. Furthermore, the best overall performance is achieved by the Ensemble COV+CORR configuration with *N*_hd_
= 50.

| | $N_{\text{hd}} = 20$ | $N_{\text{hd}} = 35$ | $N_{\text{hd}} = 50$ |
| --- | --- | --- | --- |
| ATM | **$0.52 \pm 0.10$** | $0.45 \pm 0.04$ | **$0.52 \pm 0.08$** |
| COV | **$0.67 \pm 0.08$** | $0.64 \pm 0.04$ | **$0.67 \pm 0.07$** |
| CORR | $0.51 \pm 0.04$ | $0.50 \pm 0.05$ | **$0.54 \pm 0.06$** |
| Ensemble COV+ATM | **$0.69 \pm 0.06$** | $0.66 \pm 0.04$ | $0.68 \pm 0.09$ |
| Ensemble COV+CORR | $0.72 \pm 0.07$ | $0.71 \pm 0.06$ | **$0.76 \pm 0.05$** |

